# From genome to phenome via the proteome: broad capture, antibody-based proteomics to explore disease mechanisms

**DOI:** 10.1101/2022.08.19.22278984

**Authors:** Mine Koprulu, Julia Carrasco-Zanini, Eleanor Wheeler, Sam Lockhart, Nicola D. Kerrison, Nicholas J. Wareham, Maik Pietzner, Claudia Langenberg

## Abstract

Studying the plasma proteome as the intermediate layer between the genome and the phenome has the potential to identify disease causing genes and proteins and to improve our understanding of the underlying mechanisms. Here, we conducted a *cis*-focused proteogenomic analysis of 2,923 plasma proteins measured in 1,180 individuals using novel antibody-based assays (Olink® Explore 1536 and Explore Expansion) to identify disease causing genes and proteins across the human phenome. We describe 1,553 distinct credible sets of protein quantitative trait loci (pQTL), of which 256 contained cis-pQTLs not previously reported. We identify 224 cis-pQTLs shared with 578 unique health outcomes using statistical colocalization, including, gastrin releasing peptide (GRP) as a potential therapeutic target for type 2 diabetes. We observed convergence of phenotypic consequences of cis-pQTLs and rare loss-of-function gene burden for twelve protein coding genes (e.g., *TIMD4* and low-density lipoprotein metabolism), highlighting the complementary nature of both approaches for drug target prioritization. Proteogenomic evidence also improved causal gene assignment at 40% (n=192) of overlapping GWAS loci, including *DKKL1* as the candidate causal gene for multiple sclerosis.

Our findings demonstrate the ability of broad capture, high-throughput proteomic technologies to robustly identify new gene-protein-disease links, provide mechanistic insight, and add value to existing GWASs by enabling and refining causal gene assignment.

## Introduction

Rare and common sequence variation across the genome contributes to the risk of most human diseases investigated to date (1). However, the translation of the many established and emerging genome-to-phenome links is limited by the uncertainty around the underlying causal genes. This presents a major limitation for experimental follow-up, mechanistic understanding, and use of the emerging genomic evidence in drug development. Different approaches, such as integration of tissue-specific gene expression data (2), experimentally derived functional genomic data such as ChIP-seq or ATAC-seq (3), or functional characterization of candidate variants using CRISPR screens in cellular models (4) have been used to address this gap and to identify likely causal genes at risk loci. However, complex regulatory processes take place at each stage of transcription and translation, which often leads to low correlation between transcripts and proteins, and cellular models can only approximate complex human biology. Compared to these methods, the proteogenomic approach has the advantage of focusing on the biologically active entity - the protein.

The development of broad-capture proteomic assays, targeting thousands of proteins in parallel, now enables proteogenomic approaches which can efficiently identify causal genes by systematically testing for shared genetic regulation of protein levels or function and disease susceptibility. This has catalyzed substantial advances in the identification of a) causal genes and proteins underlying established disease ‘loci’, and b) molecular ‘hubs’ that connect the genome not to one but many diseases through the encoded protein (5-18). Previous large-scale proteogenomic studies covering thousands of proteins have almost exclusively used aptamer-based assays (10, 11, 15, 16). Correlations of protein measures from aptamer versus antibody-based technologies have been shown to vary widely, and proteogenomic results are concordant for around only 65% based on around 900 overlapping proteins targets (16). To date, antibody-based proteomic assays have only been available for selected protein panels at scale (9, 14, 18), but this is changing with the availability of the Olink® Explore 1536 and Olink® Explore Expansion assays measuring ∼1,400 proteins each.

The UK Biobank Pharma Proteomics Project (UKB-PPP) project which measured ∼1,400 proteins using Olink® Explore 1536 assay in over 50,000 participants successfully demonstrated the power of scaling up by cataloguing over 10,000 mainly novel pQTLs (17). However, this study provided few insights about the translational potential of pQTLs to systematically inform candidate gene annotation at known risk loci and more importantly, to reveal novel biological roles of proteins for human health at scale. UKB-PPP and others did demonstrate that genuine and biologically relevant protein quantitative trait loci (pQTL) can be discovered in as few as hundreds of individuals (14, 17, 19), suggesting that broader proteomic coverage in even small-scale proteogenomic studies can make substantial advances to the understanding of diseases if integrated with large-scale phenomic data.

Here we generate antibody-based proteomic data using the Olink® Explore 1536 and Explore Expansion assays to capture 2,923 proteins in 1,180 individuals. We perform genetic fine-mapping at protein coding genes (±500kb) and enhance the understanding of disease mechanisms by systematically integrating cis-pQTLs with thousands of diseases and health measures to (a) refine the candidate causal gene assignment at existing disease susceptibility loci at scale and (b) identify novel disease mechanisms in phenome-wide colocalization analyses.

## Results

### Identification and fine-mapping of cis-proteogenomic signals for 2,923 protein targets

We adopted a Bayesian fine-mapping strategy (20) to identify proximal acting genetic variants (cis-pQTLs, ±500kb around the protein coding gene) that were associated with plasma abundance of 2,923 proteins measured in 1,180 participants of the EPIC-Norfolk cohort (21) (**Supplementary Table 1**). We identified a total of 1,553 independent credible sets for 914 unique protein targets for which sentinel variants reached genome-wide significance (p<5×10^−8^) when modelled jointly at each protein coding locus (**Fig. 1A, Supplementary Table 2**). The number of independent credible sets for each protein target ranged between one and eight (mean=1.64, IQR=1-2), illustrating wide-spread allelic heterogeneity at protein coding loci. We observed a high replication rate (89.9%, 910 out of 1,013) for credible sets of 590 protein targets overlapping with the UKBB-PPP effort. Conversely, we identified 4.5% of the 20,540 reported signals in as few as 1,200 participants that were reported based on more than 35,000 participants of the UKB-PPP. A total of 256 (16.5%) credible sets contained cis-pQTLs not previously reported, including 131 proteins that have not been measured by previous platforms (**Fig. 1B**) (5-18). Notably 125 signals were for 101 previously targeted proteins, the majority of which (n=92 proteins) have been targeted using non-antibody-based technologies in samples sizes up to 30 times larger than ours (10, 11, 15, 16) (**Fig. 1B**).

**Figure 1:**
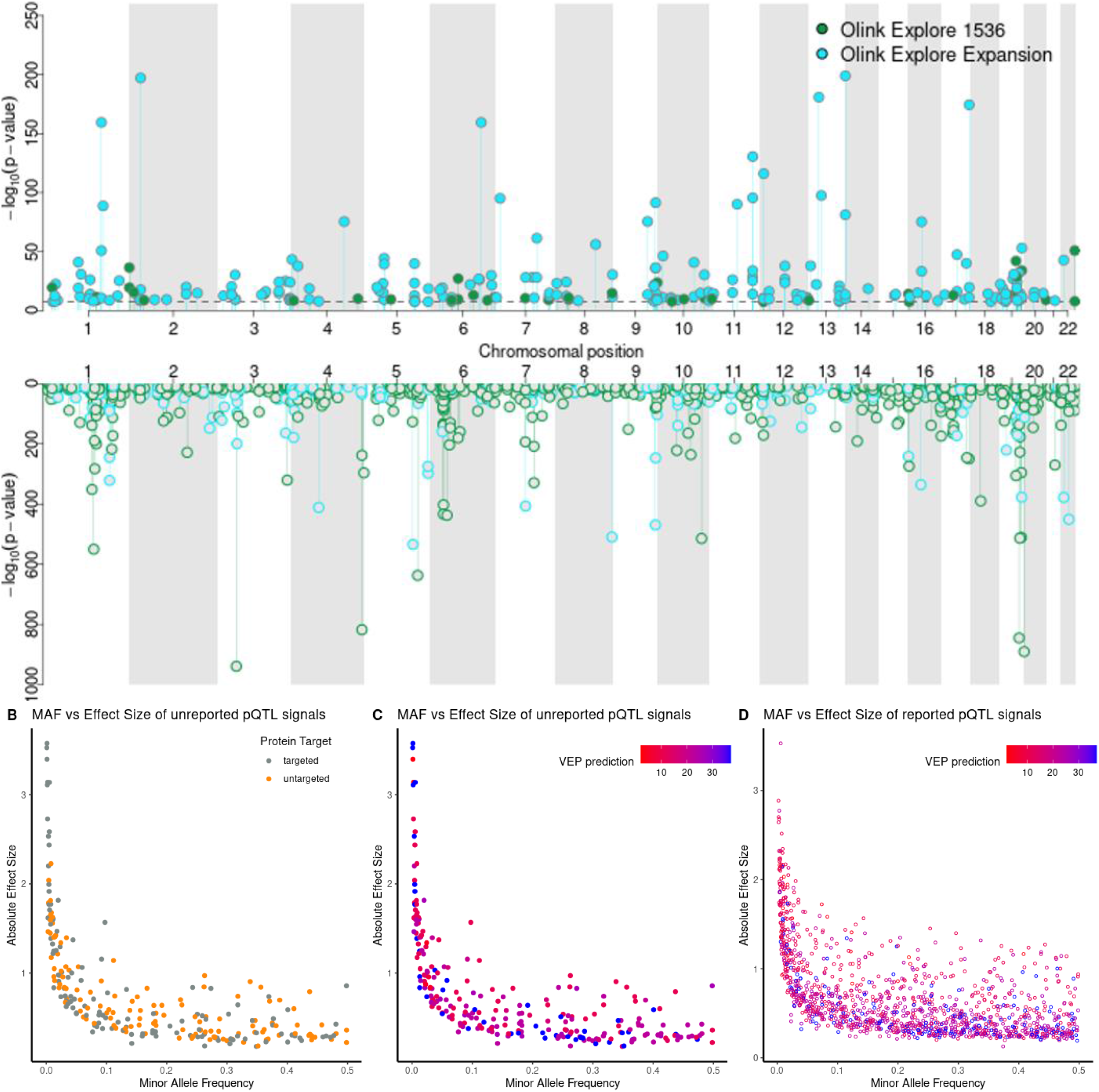
Genetic regulation of 2,923 proteins measured by the Olink Explore 1536 and Olink Explore Expansion platforms in 1,180 individuals. Previously unreported and reported pQTLs are represented with a filled and hollow circle, respectively. Only the variants which are genome-wide significant (p-value<5×10^−8^) in the joint model (see Methods) are presented. **A. Miami plot representing the independent lead cis-pQTLs identified through Bayesian fine-mapping for 914 unique proteins**. Shown are p-values from a linear regression model modelling all identified credible set variants for a given protein target jointly. *Top*: Lead cis-pQTL signals unreported to date. *Bottom*: Lead cis-pQTL signals which were in linkage disequilibrium (LD; r^2^>0.5) with a previously reported pQTL. **B. Minor allele frequency vs effect size of unreported pQTL signals, coloured by whether the protein has previously been targeted**. Unreported pQTL signals for a previously targeted protein are coloured grey and those for a previously untargeted protein are coloured orange. **C. Minor allele frequency vs effect size of unreported pQTL signals, coloured by most severe variant consequence prediction**. The colour coding represents the most severe Variant Effect Predictor (22) consequence of the lead cis-pQTL, or variants in LD (r^2^>0.6) within the protein encoding gene. The most severe consequence is coloured red (Ensembl consequence rank = 1) and the least severe consequence is coloured blue (Ensembl consequence rank = 37). **D. Minor allele frequency vs effect size of reported pQTL signals, coloured by most severe variant prediction**. The colour coding represents the most severe Variant Effect Predictor (22) consequence of the lead cis-pQTL, or variants in LD (r^2^>0.6) with the lead cis-pQTL within the protein encoding gene. The most severe consequence is coloured red (Ensembl consequence rank = 1) and the least severe consequence is coloured blue (Ensembl consequence rank = 37).

Effect size and minor allele frequency distributions of unreported cis-pQTLs were comparable to the 1,297 (83.5%) successfully replicated cis-pQTLs (5-18) (**Supplementary Table 2**), illustrating that complementary proteomic technologies can still identify genetic variants that would have been anticipated to be seen in previous studies (**Fig. 1A, Fig. 1D**).

We observed a strong inverse relationship between the absolute effect sizes of cis-pQTLs and the log_10_-transformed frequencies of their minor alleles (r=-0.78; p<1×10^−300^), likely due to the more severe predicted consequences of rarer alleles, such as stop-gain mutations (**Fig. 1B-C**). We also report 482 cis-pQTLs with a minor allele frequency (MAF) above 5% with large absolute effect sizes (range 0.5-1.72 s.d. per allele), suggesting strong genetic control of the associated proteins. Of these, less than half (35.9%) were protein altering variants themselves or were in strong linkage disequilibrium (LD; r^2^>0.6) with one, potentially affecting the binding affinity of antibodies. Proteins with at least one significant cis-pQTL were enriched for characteristics of secreted proteins, like the presence of disulfide-bonds (odds ratio: 4.47; p-value=3.0×10^−74^) or glycosylation sites (odds ratio: 2.22; p-value=1.1×10^−13^), but depleted of sites for posttranslational modifications that are important for intracellular signaling, like phosphorylation (odds ratio: 0.42; p-value=5.5×10^−14^) or ubiquitination (odds ratio: 0.30; p-value=4.2×10^−11^).

Finally, for more than half of the protein targets (n=532) with at least one pQTL, we observed strong evidence of colocalization (PP>80%) between a cis-pQTL and the corresponding gene expression QTL (eQTL) signal in at least one out of 49 tissues of the GTEx resource (**Supplementary Table 3**). These results suggest altered expression of protein coding genes in one or multiple tissues as the major source for cis associations observed with plasma protein levels.

### From genome to phenome via the proteome

The genome is linked to the phenome via the proteome and the translational potential of pQTLs is due to their ability to link insights about the genetic regulation of protein levels and function to diseases (15). We identified 1,110 robust protein – phenotype pairs (**Fig. 2**; posterior probability [PP] > 80% of a shared genetic signal) comprising 224 protein targets for 575 unique traits by systematically testing for a shared genetic architecture at protein coding loci (±500kb) across the phenome (see **Methods**; **Supplementary Table 4)**. This included well-described examples, such as UMOD and kidney disease or established drug targets like PCSK9 and LDL-cholesterol, but importantly 93 protein targets connected with at least one phenotype that have been missed by previous aptamer-based efforts.

**Figure 2:**
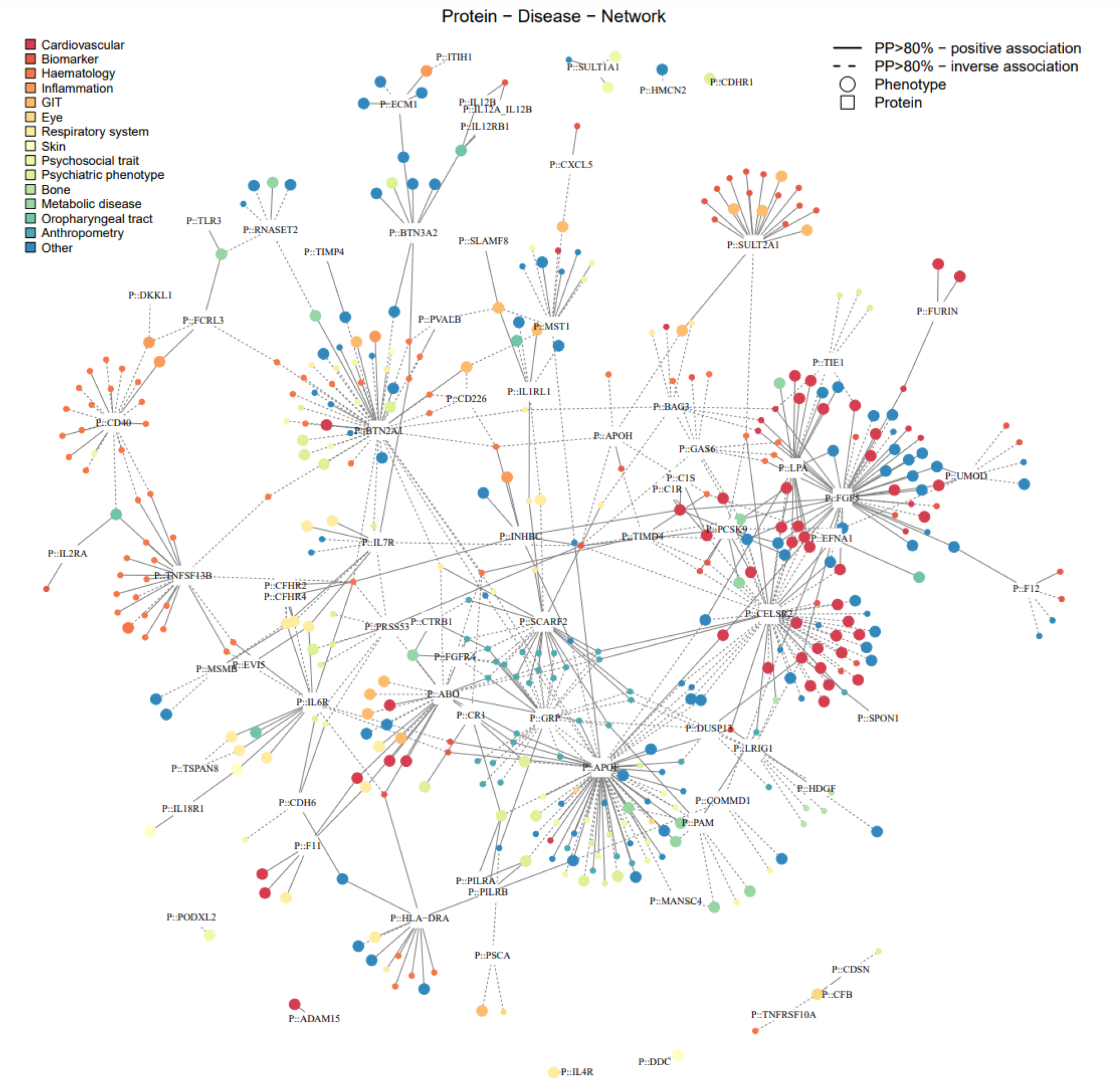
Protein – disease network. Results from phenome-wide colocalization at protein coding loci (±500kb) are shown. For simplicity, only proteins with at least one binary outcome (i.e., mainly diseases) association are included. Proteins are presented with a square, binary outcomes are presented with large circles, and continuous outcomes are presented with small circles. The colour for the circles present the trait category. Edges between proteins and phenotypes represent strong evidence for a shared genetic signal (PP>80% and LD between regional sentinel variants >0.8). Effect directions are indicated by the line type (solid = higher protein abundance, increased risk, dashed = higher protein abundance, reduced risk) and derived based on the lead cis-pQTL at the corresponding locus. The full list of colocalization results can be found in Supplementary Table 4. Abbreviations: GIT, gastrointestinal tract.

One of the examples is gastrin releasing peptide (GRP, encoded by *GRP*), for which we observed strong evidence of colocalization (posterior probability [PP] =82.5%) between plasma levels and type 2 diabetes (T2D) risk at an established GWAS locus (18q21) for which different genes had been prioritized, including *SEC11C, GRP*, and *MC4R* (23-25). The GRP-increasing G-allele of the lead cis-pQTL (rs1517035; MAF=0.18) was associated with a reduced risk for T2D (odds ratio=0.96, p-value=7.8×10^−10^). GRP is a neuropeptide named for its ability to stimulate secretion of the gastric acid secretagogue, gastrin, in the stomach (26, 27), but it is likely involved in other metabolic pathways. We obtained strong evidence that GRP likely mediates T2D risk via an effect on overall obesity, based on the convergence of evidence from mice studies, human trials, and human genetic data. We established a shared genetic signal between plasma GRP, body mass index and fat, and T2D risk using multi-trait colocalization with coherent effect directions (**Fig. 3**). GRP induces satiety in mice via its cognate GRP receptor (*Grpr)* (28, 29). Further, mice lacking *Grpr* show impaired glucose tolerance after gastric glucose administration (30) and gain excess body weight under *ad libidum* conditions (29). These observations have been corroborated by human trials, in which treatment with human recombinant GRP (hrGRP) led to weight loss through reduced food intake (31). In summary, our results motivate investigations into hrGRP for appetite control and body weight lowering to possibly assist in T2D management and remission, an approach similar to recently implemented treatment strategies targeting incretins, like GLP-1, and associated receptors, with preliminary evidence of an additive effect in rats (32).

**Figure 3:**
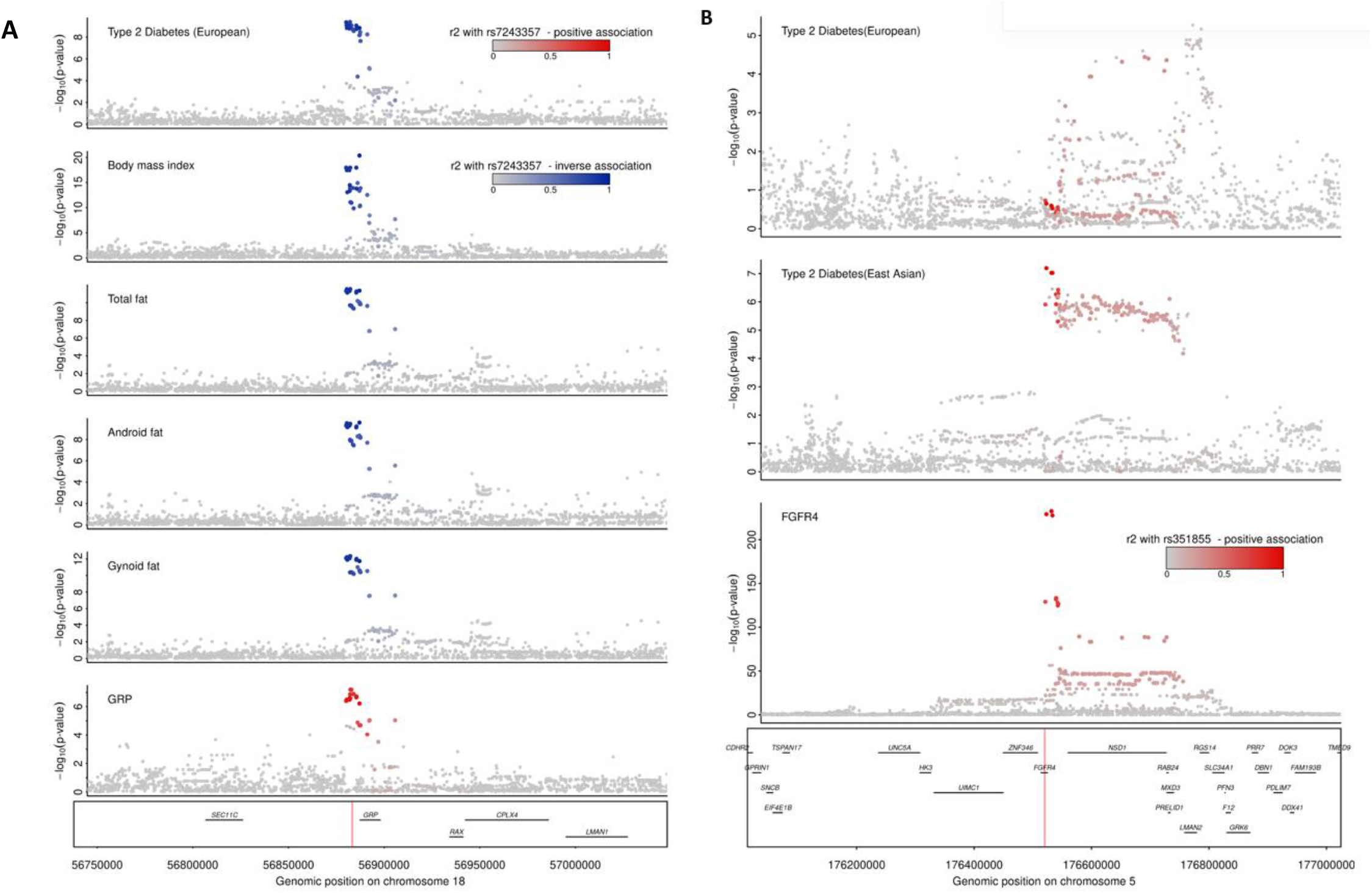
Stacked regional association plots for the multi-trait colocalization. **A. Stacked regional association plots for the multi-trait colocalization of the GRP cis-pQTL with gynoid fat, android fat, total body fat, body mass index and type 2 diabetes.** The top candidate SNP highlighted by multi-trait colocalization (rs7243357) and lead cis-pQTL for GRP (rs1517035) are in strong LD (r2=0.8). Gynoid fat, android fat and total body fat phenotypes are based on UK Biobank and were analysed in-house using BOLT-LMM (33). **B. Stacked regional association plot the multi-trait colocalization of the FGFR4 cis-pQTL with type 2 diabetes in East Asian populations**. Red colouring represents a positive effect direction in reference to the protein increasing allele for GRP whereas blue represent an inverse association. The hue of the colour represents the strength of r^2^ representing the LD structure, as indicated on the legend. European Type 2 diabetes summary statistics were obtained from dbGAP Million Veteran Program (MVP) European subset (n_cases_= 148,726, n_controls_= 965,732) (34). East Asian Type 2 diabetes summary statistics were obtained from Mahajan et al (2022) (n_cases_= 56,268, n_controls_= 227,155) (25). The body mass index summary statistics were obtained from Pulit et al. (2019) (n=806,834) (35).

Several T2D loci have been reported to be specific to certain ancestries (23-25). In the absence of strong differences in allele frequency, such ancestry specific effects could be caused by a variety of different factors, including environmental factors such as dietary intake. We obtained robust evidence that *FGFR4* is the candidate causal gene at the East Asian-specific *FGFR4*-*NSD1* locus supported by a high posterior probability (PP=97%) for a shared genetic signal with plasma levels of the gene product fibroblast growth factor receptor 4 (FGFR4) and trans-ancestral conserved LD between regional sentinel variants (r^2^>0.96; **Fig. 3**). The protein-increasing A-allele of the lead cis-pQTL (rs351855, beta= 1.01, p-value=9.8×10^−234^, EAF_European_=0.30, EAF_EastAsian_=0.46) was associated with an increased risk for T2D (beta=0.05, p-value=1.1×10^−7^). Candidate gene studies have implicated rs351855 (p.G388R) in cancer susceptibility (36-38), and subsequent mechanistic studies showed a gain of function of the mutant FGFR4 by binding transducer and activator of transcription 3 (STAT3) (39). While we found no evidence for an association to cancer, there are different studies that support our observation of FGFR4 in T2D-related pathways including hepatic glucose, bile, and lipid metabolism, and possibly insulin signaling in a diet-dependent manner (40-43). Briefly, *Fgfr4*^*-/-*^ mice fed a normal chow diet exhibit insulin resistance and impaired glucose tolerance compared to wild-type controls, however, this difference is not observed in high-fat diet fed mice. A similar masked genetic effect is seen with the mutant protein in mice and small observational studies in humans (44). The ability of diet to obscure genetic effects may explain the ancestral-specific effect in the absence of strong differences in allele frequencies, with high-fat diet conditions being substantially more common in Western-style countries of predominantly European ancestry compared to East Asia (45), in particular Japan, in line with Biobank Japan (p-value_T2D_<7.6×10^−11^) being the largest contributing population to the East Asian T2D meta-analysis (25).

### Proteogenomic guided annotation of genes at loci reported for diseases and traits related to human health

Annotation of the candidate causal genes at disease susceptibility loci is the major bottleneck in the translation of GWAS into biological and possibly clinical insights (46). We exploited the genomic proximity between cis-pQTLs and the protein coding gene for gene annotation by systematically overlapping identified credible sets in this study with reported risk loci (p<5×10^−8^) from the GWAS catalog (downloaded on 23/03/2022; (1)). We identified 480 credible sets targeting 395 unique proteins (43.2% of all, 914 unique protein targets) for which the lead cis-pQTL or a proxy (r^2^>0.8) had been reported for one or more of 5,391 collated traits in the GWAS catalog (**Fig. 4 and Supplemental Tab. 5, see Methods**). For 40% (n=192) of those, we prioritized a gene that was different from the one originally reported, of which 50% (n=96) were not the gene nearest to the GWAS sentinel variant. We further refined a longer list of putative causal genes to a single one for an additional 31 cis-regions (6.5%). These results exemplify the unique potential of cis-pQTLs for gene annotation of loci reported across diseases and traits related to human health (**Fig. 4 and Supplemental Tab. 5**), with one example outlined in more detail below.

**Figure 4:**
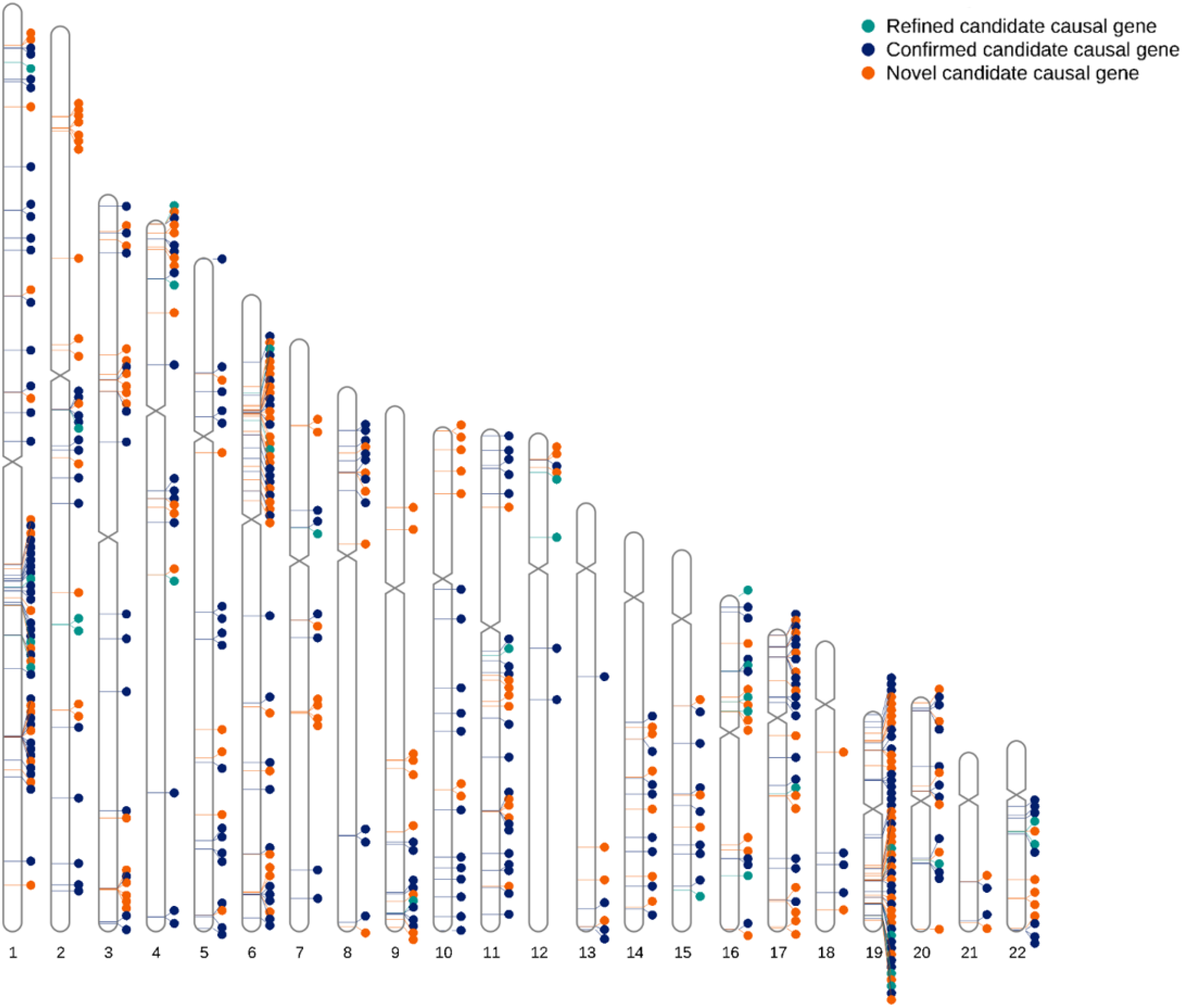
Candidate causal gene assignment at reported GWAS loci using pQTLs. The overlap between existing GWAS risk loci and pQTL loci (n=480) are marked on the human chromosome karyotypes (chromosomes 1-22). The locus is coloured orange if the pQTL provides a novel candidate causal gene assignment for one or more traits, light blue if it refines a candidate causal gene from a longer list of reported or closest genes, and dark blue if it confirms the candidate causal gene assignment provided by the GWAS.

Multiple Sclerosis (MS) is an autoimmune, inflammatory, and neurodegenerative disease of the central nervous system that is caused by both genetic and strong environmental factors (47). A strong signal at 19q13.33 is one of 233 reported GWAS loci (48). Several variants in high LD (r^2^>0.9) reported for this locus have been linked to different candidate causal genes, including *DKKL1, CD37*, and *SLC6A16* which was the most recently annotated gene based on a sophisticated ensemble of methods (48). We identify a shared genetic signal (PP=96.1%, **Supplemental Fig. 1**) between dickkopf like acrosomal protein 1 (DKKL1), encoded by *DKKL1*, and MS at this locus, led by a cis-pQTL (rs2288480; MAF=0.25) in high LD (r^2^=0.97) with the lead MS variant (rs1465697; MAF=0.33, OR=1.09, p-value=3×10^−18^). We note that the lead cis-pQTL was also in LD (r^2^=0.97) with a recently identified variant at the same locus for systemic lupus erythematosus (SLE) among East Asians (49).

The lead cis-pQTL is in strong LD (r^2^>0.8) with a cluster of three common missense variants (rs2288481, rs2303759, and rs1054770) that might impair protein function or processing. Little is known about the biological role of DKKL1 in general, but a non-essential role in spermatogenesis has been described (50). However, a link towards MS and/or SLE might be conceivable via a possible role of DKKL1 in adaptive immunity and hence the inflammatory component of MS. Briefly, *DKKL1* expression is enriched among memory B-cells (51) and an independent secondary cis-pQTL (rs66532151, MAF=22.4%) for DKKL1 tagged (r^2^>0.96) a cluster of variants associated with different characteristics of CD20^+^ memory B-cells (52). This cis-pQTL (rs66532151) was associated with MS at p-value=3.4×10^−7^, providing late genetic evidence for depletion of B-cells being one of the most effective treatments for MS, a therapeutic strategy that originally emerged from clinical and neuropathological studies (53, 54). Further follow-up studies are needed to clarify a possible role of DKKL1 in immune cells and whether DKKL1 may play a role in B-cell hyperactivity observed in MS (53).

Multiple independent genetic variants associated with the same protein target at the same locus, so-called allelic heterogeneity, provides the highest confidence in gene assignment but can also highlight differential biological roles for the same protein. We observed 73 such protein targets with two or more credible sets including distinct GWAS variants for related and unrelated traits. For example, we discovered three distinct credible sets for plasma levels of interleukin 34 (IL-34) at 16q22.1. Two contained independent (r^2^=0.07) lead cis-pQTL variants with distinct structural consequences on the protein, that were also associated with two distinct outcomes – Alzheimer’s disease (55) and childhood obesity (56) (**Fig. 5**). rs4985556 is associated with increased risk for Alzheimer’s disease (MAF=12.2%; beta=0.07, p-value<2.3×10^−8^) and introduces a premature stop (p.Tyr213Ter), truncating the protein and likely affecting dimerization and possibly secretion. rs8046424 (alternate allele: C; r2=0.96 with lead sentinel childhood obesity variant rs4985555) is associated with reduced childhood obesity (MAF = 48.2%; beta=-0.008, p-value<4.4×10^−9^). It is a missense variant (p.Glu123Gln) of moderate consequence (CADD score 11.6) that maps to a binding domain of the cognate IL-34 receptor CSF-1R (57). Therefore, both variants will likely strongly (rs4985556) or moderately (rs8046424) attenuate signaling via CSF-1R, which has been shown to drive cerebrovascular pathologies that are common in Alzheimer’s disease (58). While the gradient of structural consequences translates into a graded effect on Alzheimer’s disease (rs8046424, beta=0.03, p<3.6×10^−4^), the absence of any effect of the more detrimental variant (rs4985556) on childhood obesity (beta=-0.001, p=0.59) might point to a different, yet to be defined, pathway.

**Figure 5:**
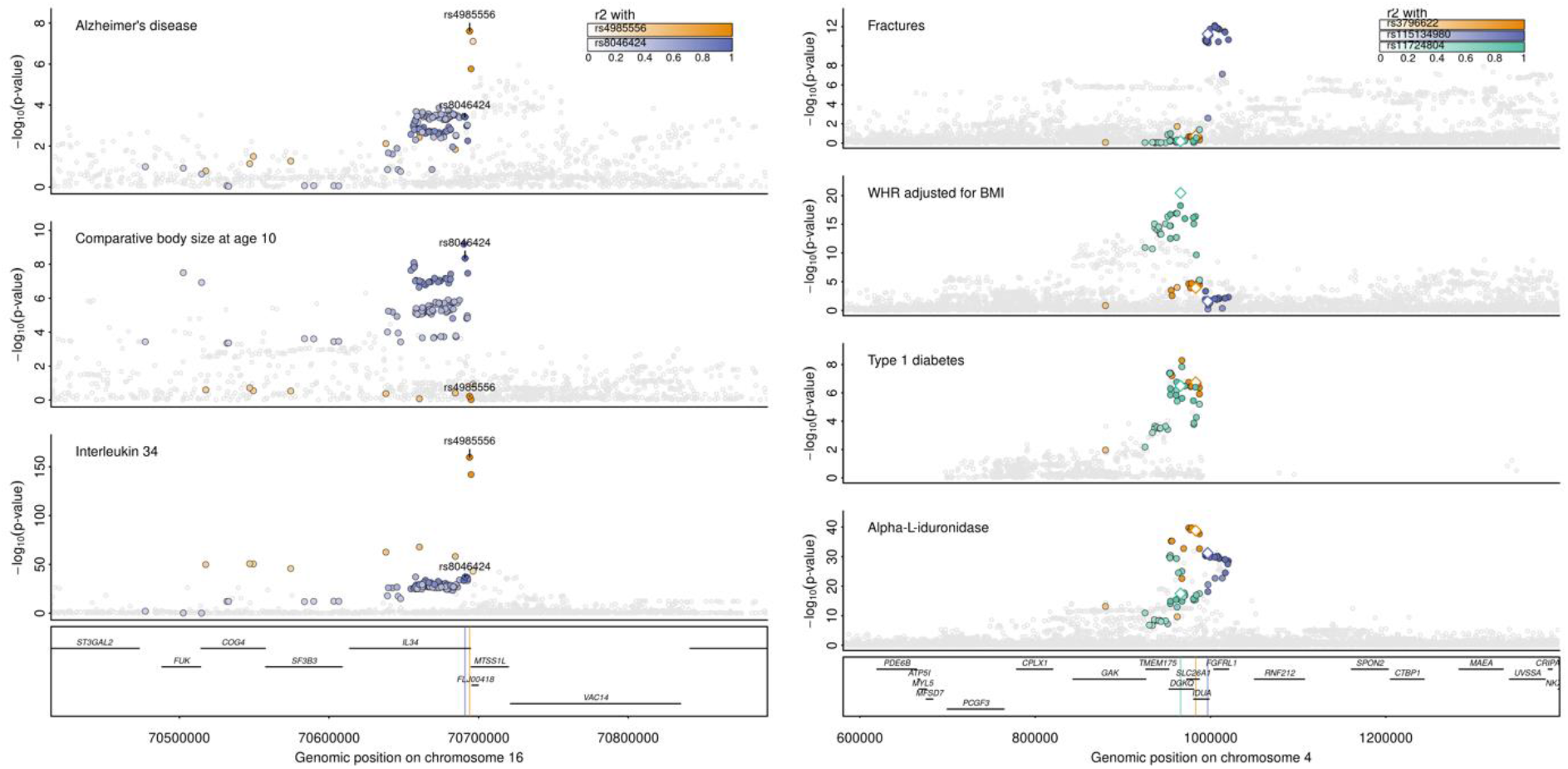
Allelic heterogeneity at protein coding loci translates into distinct phenotypic consequences at *IL34* and *IDUA*. *Left* Regional association plots centered around *IL34* (±200kb) for plasma interleukin 34 levels, comparative body size at age 10 (56), and Alzheimer’s disease (55). Shown are association statistics (p-values) from genome-wide association analysis. Single genetic variants were coloured based on LD with two distinct cis-pQTLs (rs4985556 – orange; rs8046424 – purple). *Right* Regional associations plots centered around IDUA (±400kb) for plasma alpha-L-iduronidase levels, type 1 diabetes (62), waist-to-hip ratio (WHR) adjusted for body mass index (BMI) (60), and risk of fractures (59). Shown are association statistics (p-values) from genome-wide association analysis. Single genetic variants were coloured based on LD with three distinct cis-pQTLs (rs3796522 – orange; rs115134980 – purple; rs11724804 – green). Lead cis-pQTLs are highlighted by hollow diamonds.

We observed a similar segregation of phenotypes across distinct cis-pQTLs for alpha-L-iduronidase encoded at *IDUA*. Briefly, three out of four detected credible sets contained GWAS risk loci or strong proxies (r^2^>0.8) for fractures (59) (rs115134980; MAF=16.1%; beta=-0.06, p-value=7.4×10^−12^), waist-to-hip ratio adjusted for BMI (60) and inflammatory diseases (61) (rs11724804; MAF=44.7%; beta=-0.017, p-value<7.6×10^−21^), as well as type 1 diabetes (62) (rs3796622; MAF=35.2%; beta=-0.07, p-value<1.7×10^−7^) (**Fig. 5**). Alpha-L-iduronidase is essential for the breakdown of glycosaminoglycans within lysosomes and numerous rare pathogenic variants within *IDUA* are known to cause accumulation of glycosaminoglycans in lysosomes (mucopolysaccharidosis type I [MPS-1]). Patients present with a wide spectrum of complications, such as skeletal deformities or organomegaly, that has been attributed to the variable impact of mutations on enzyme activity, with nonsense mutations causing most severe diseases (Hurler syndrome) (63). While skeletal abnormalities in rare disease patients may relate to bone phenotypes seen for the common cis-pQTL, there are no reports for an elevated risk for inflammatory or autoimmune disease among MPS-1 patients or other evidence from rare variant analysis. Tissue-dependent effects of common variants might be one explanation for the different phenotypes linked to distinct cis-pQTLs for alpha-L-iduronidase.

### Phenotypic convergence of rare variant burden and common cis-pQTLs for protein coding genes

Much effort and funding has been invested into biobank-scale whole-exome sequencing studies (ExWAS) to identify rare deleterious genetic variants and novel disease candidate genes for the development of treatment strategies (64, 65). However, it is unknown how efforts focusing on the rare deleterious end of gene (and protein) dysfunction relate to less extreme alterations of protein levels or function. To explore whether evidence from ExWAS and our cis-based phenome-wide colocalization analyses converge for disease-linked genes, we systematically integrated our results with those from a recent ExWAS among ∼450,000 UK Biobank participants across almost 4,000 phenotypes (64).

Among 2,939 protein coding genes covered by the Olink Explore 1536 and Explore Expansion platforms, 40 (1.3%) showed evidence for phenotypic associations with a rare variant gene-burden and statistical colocalization with a cis-pQTL, whereas 281 and 184 protein coding genes were linked to phenotypes through ExWAS or cis-pQTLs only, respectively (**Fig. 6**). Out of the 40 overlapping genes, we observed phenotypic convergence for only 12 genes across 21 phenotypes following manual review to harmonize phenotype definitions (**Supplementary Tab. 6**). These results clearly exemplify the complementary nature of both approaches and the unique ability of bespoke proteogenomic experiments to prioritize disease mediators and hence putative therapeutic targets.

**Figure 6:**
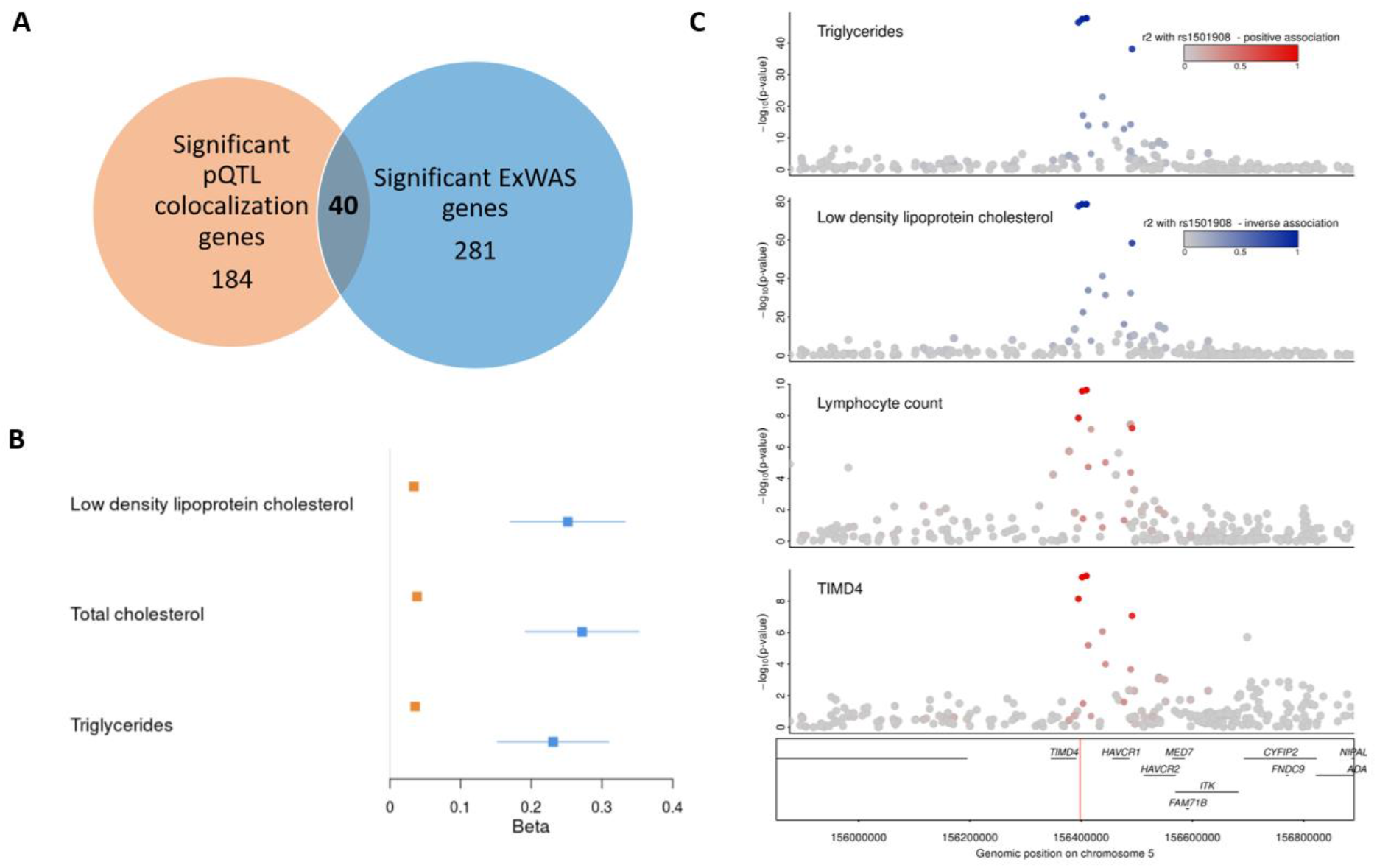
Phenotypic convergence of rare variant burden and common cis-pQTLs for protein coding genes and TIMD4 as an example. A. Venn diagram showing the number of genes with a significant rare variant gene burden association (p<1E-06) with at least one trait (64) in blue and the number of genes with a significant pQTL colocalization (PP>80%) with at least one trait in orange. All 2,939 unique genes covered by Olink Explore 1536 and Explore Expansion assays were investigated. **B. Forest plot comparing the effect size estimates between TIMD4 cis-pQTL (rs58198139) and rare TIMD4 loss of function (LoF) gene-burden results (variant group: missense and loss of function variants with a minor allele frequency < 1%) for low density lipoprotein cholesterol, total cholesterol and triglyceride levels**. Rare TIMD4 loss of function (LoF) gene-burden results are shown in blue and TIMD4 cis-pQTL associations are shown in orange. C. **Stacked regional plot of the multi-trait colocalization of TIMD4 cis-pQTL with lymphocyte count, low density lipoprotein cholesterol, and triglycerides**. Red colouring represents a positive effect direction with protein increasing allele with TIMD4 whereas blue represent an inverse association. The hue of the colour represents the strength of r^2^ representing the LD structure, as indicated on the legend.

Convergence of phenotypic consequences from rare gene burden and common cis-pQTLs not only provides compelling evidence for causal gene assignment but can establish dose-response relationships that are an essential prerequisite for genetically informed drug discovery (66). We observed such a dose-response relationship between putative functional consequences for T-cell immunoglobulin and mucin domain containing 4 (TIMD4) and LDL-cholesterol as well as total triglyceride, but not HDL-cholesterol levels in blood (**Fig. 6C)**. The protein-decreasing T-allele of the lead cis-pQTL (rs58198139) was associated with moderate effects on LDL-cholesterol in UK Biobank (MAF=0.26; beta_LDL_=0.03, p-value_LDL_=7×10^−44^), likely mediated by altered protein expression, while the cumulative burden of rare loss-of-function variants was associated with substantially higher LDL levels (beta_LDL_= 0.25, p-value_LDL_= 1.51×10^−9^, variant mask: predicted loss of function and deleterious missense variants with MAF<1%), in line with this locus being one of the earliest discovered loci for polygenic dyslipidemia but with few functional insights gained since (67). TIMD4 is best known for its role in tissue-dependent macrophage efferocytosis of apoptotic cells (68, 69) but does also participate in T-cell activation and recruitment (70). Accordingly, *Timd4*^*-/-*^ mice show impaired macrophage phagocytosis and increased lymphocyte cell counts (71), an observation recapitulated by our phenome-wide colocalization analyses identifying an inverse association for the protein-decreasing T-allele for lymphocyte counts (beta=1.02, p-value=1.4×10^−12^) with high certainty (PP = 97.5%). Circulating leucocytes and resident M2 macrophages can take up cholesterol from circulating LDL particles and sequestered lipoproteins in the vasculature, but classical pathways, like the LDL-receptor mediated uptake, were shown, at least in mice, to have no substantial effect on plasma LDL-cholesterol levels (72). In contrast, more recent work demonstrated the ability of TIMD4^+^ adipose tissue macrophages to significantly contribute to the regulation of post-prandial HDL-cholesterol levels in mice (73). While there were was no difference in triglycerides or non-HDL cholesterol following TIMD4 blockade, TIMD4 blockade inhibited LDL-induced lysosomal activity *in vitro*, suggesting a role for TIMD4 in peripheral LDL cholesterol processing. These findings provide evidence of a role for TIMD4 in the regulation of systemic lipoprotein metabolism, and taken together with our proteogenomic findings, provide a compelling rationale to explore the role of TIMD4^+^ macrophages in systemic LDL cholesterol metabolism. In general, increased uptake of modified LDL-cholesterol particles by resident macrophages contributes to atherosclerotic foam cell formation, a major cardiovascular risk factor (74). We observed no conclusive evidence that either the cis-pQTL (protein decreasing T-allele; odds ratio [95% CI] = 1.04 [1.02-1.07], p-value=0.002) or the cumulative burden of the loss of function variation in the gene (odds ratio [95% CI] = 1.3 [0.90-1.88], p-value=0.16) were associated with coronary artery disease (CAD; the currently most powered GWAS for atherosclerotic consequences). However, our findings urge further investigations into the functional role of TIMD4 in immune cell-mediated LDL-cholesterol turnover and foam cell formation. Although the extent to which this mechanism can contribute to addressing CAD burden is currently unclear, blocking of TIMD4 using monoclonal antibodies increased atherosclerotic lesion size in *Ldlr*^*-/-*^ mice although in a possibly cholesterol-independent manner (75).

## Discussion

Proteogenomic approaches have the potential to establish a direct link from rare and common variation in or closeby protein-encoding genes to human health via the protein product (5-18). Despite recent advances and early successes, the field is still in its infancy with respect to the scale and protein capture, with existing broad-capture technologies currently targeting less than a third of all proteins encoded in the human genome (5-18), not capturing posttranslational modifications, or providing absolute protein quantification.

Here we identified more than 200 novel cis-pQTLs that have not been reported so far, even in studies 30-times larger than ours using non-antibody-based technologies, by capitalizing on recent assay developments. The fact that we identified hundreds of cis-pQTLs for proteins that have been investigated in studies much larger than ours might be best explained by the need to develop further orthogonal methods to measure protein targets as we have outlined previously (14).

We demonstrate that systematic application of cis-pQTLs to large-scale genetic studies of human diseases can 1) guide causal gene annotation at GWAS loci (e.g., *DKKL1* for multiple sclerosis), 2) identify pathways that link genes to diseases guided by a protein-phenotype network, and 3) complement gene-burden testing of rare variants to discover novel biology. We highlight specific examples in more detail and share a large number of high-confidence protein–phenotype associations that provide a direct guide for functional follow-up and future investigations of variant protein with disease relevance about which little is known to date.

The vast majority (∼90%) of genetic variants identified in GWASs reside in non-coding regions of the genome (76), creating a challenge for variant-to-function annotation. In line with previous studies, we demonstrate the efficiency and ability of cis-pQTLs to prioritize causal candidate genes including reassignments at 40% of overlapping loci. In contrast to other annotation approaches, the particular value of the integration of proteogenomic studies lies in the instrumentalization of the likely biological effector molecules. Studies using proteomic profiling in disease relevant tissues or single cell-types are needed to further elucidate the mechanisms underlying the many thousands of unassigned GWAS loci.

This study is a powerful demonstrating that even moderately sized proteomic studies can result in the identification of novel biology when combined with bespoke analysis pipelines designed for the identification of cis-pQTL and systematic integration and follow-up of disease GWAS summary statistics. Eventually multiple different technologies will be needed at scale to capture not only proteins of interest but also the vast spectrum of proteoforms with possible distinct phenotypic consequences (14). This prediction is supported by power calculations of the UKB-PPP (17), but also based on the observation that our study identified cis-pQTLs for genes that are under less evolutionary constraint as indicated by higher observed/expected scores for missense (+0.15; p-value=5.0×10^−50^) and loss-of-function (+0.22; p-value=7.5×10^−51^) variation in gnomAD (77). This observation is in line with recent findings among eQTL studies (78).

We observed convergence of gene–phenotype associations between ExWAS and our proteogenomic approach only at a small number of genes. Gene identification with overlapping or converging evidence, as shown for *TIMD4*, provides high confidence about the underlying causal gene, while the incomplete overlap clearly indicates the complementary nature of both approaches for drug target prioritization. An important distinction between both approaches, beyond the different genetic variants covered, is the ability of proteogenomics to emulate protein variation across the whole spectrum of abundance and in some cases function, and not only putative loss-of-function (rarely gain of function), which might explain differences seen in phenotypic consequences between both approaches. In addition, in terms of practicality, integration of pQTLs into colocalization and GWAS loci annotation enabled us to uncover unreported disease biology with a small sample size of 1,180 individuals, whereas substantially larger sample sizes, even millions of individuals, are needed to reach enough power to detect rare variant associations in ExWAS studies for disease endpoints (79).

Our study has some limitations that need to be considered. Affinity-based reagents allow for the quantification of protein abundance but are inherently limited to quantify the level of activity, although a general correspondence between the two can be assumed. This limits insights about the role of protein targets using a proteogenomic approach. Further, numerous posttranslational modifications can change the function and abundance of proteins but are currently not distinguishable using affinity reagents at scale. We deliberately decided to restrict genetic analysis of protein targets to the corresponding protein coding regions (±500kb) for two reasons: 1) the high biological prior to identify genetic variants directly linked to protein function/abundance, and 2) to increase power for statistical analysis by limiting the multiple testing burden. Larger studies are needed to explore the spectrum of trans-pQTLs that have generally smaller effect sizes but can identify protein interaction partners and facilitate systematic dose-response analysis in the two-sample Mendelian randomization frameworks.

In summary, we demonstrate the clear potential of broad-capture proteogenomic studies to identify novel biological pathways that link protein-encoding genes to human diseases. Systematic integration of human genomic with proteomic and phenomic data enables such investigation even in relatively moderately sized studies and can help to prioritize targets and indications for the development of safe and effective therapeutic interventions.

## Materials and Methods

### Study participants

We measured protein levels among 1,180 participants of the European Prospective Investigation into Cancer (EPIC)-Norfolk study, a cohort of 25,639 middle-aged, individuals from the general population of Norfolk, a county in Eastern England which is a component of EPIC (21). The study was approved by the Norfolk Research Ethics Committee (ref. 05/Q0101/191) and all participants gave their informed written consent before entering the study. Information on lifestyle factors and medical history was obtained from questionnaires as reported previously (21). We selected a random sub-cohort of 771 participants and a set of 429 participants with selected incident events during follow-up for cost-efficient proteomic profiling.

### Proteomic profiling

We used serum samples from the baseline assessment (1993 - 1997) that had been stored in liquid nitrogen for proteomic profiling using the Olink Explore 1536 and Explore Expansion platforms targeting 2925 unique proteins by 2943 assays, of which 2923 unique proteins mapped to a protein encoding locus in genome assembly GRCh37. Details regarding the assay have been have been described in detail (80). Briefly, proteins are targeted by two separate unique antibodies, each of which are labelled with complementary single stranded oligonucleotides (proximity extension assays (81)). These proximity extension assays hybridization occurs subsequent to the binding of antibody pairs with complementary oligonucleotides which can be quantified using next generation sequencing (NGS). NGS read-outs undergo quality control procedures where internal (incubation, extension and amplification controls) and external (negative, plate and sample controls) controls are included. Normalized protein expression (NPX) units are generated by normalization to the extension control and further normalization to the plate control and reported on a log2 scale. We excluded 3 samples as they were shown to be extreme outliers using principal component analysis from their entire proteomic profiles. For downstream genetic analysis (fine-mapping and region-based association analysis) we first rank-inverse normal transformed NPX-values and corrected for age, sex, and the first ten genetic principal components using linear regression models. The residuals of this analysis were used throughout the study.

### Genotyping

EPIC-Norfolk samples (n=21,448) were genotyped on the Affymetrix UK Biobank Axiom array chip by Cambridge Genomic Services, University of Cambridge, UK. Sample and variant QC followed the Affymetrix Best Practices guidelines. Samples were excluded based on DishQC < 0.82 (fluorescence signal contrast), call-rate <97%, heterozygosity outliers and sex discordance checks. Variants were excluded if call-rate <95% or HWE<=1e-6. Monomorphic variants and those with cluster problems detected using Affymetrix SNPolisher were excluded. Genotype imputation was performed using two different reference panels, the Haplotype Reference Consortium (HRC) (release 1) reference panel and the combined UK10K+1000 Genomes Phase 3 reference panel. After pre-imputation QC, 21,044 samples remained for imputation. All SNPs imputed using the HRC reference panel were included, and additional variants imputed using only the UK10K+1000 Genomes reference panel were added to create a combined imputed set. Variants with imputation quality INFO < 0.4 or MAF of < < 0.0001 were excluded. All positions are on genome assembly GRCh37. After excluding ancestry outliers, individuals without a high-quality proteomic profile for each panel and pruning the sample set for related individuals, 1,180 and 1,178 individuals were included in proteogenomic analyses for Olink Explore 1536 and Explore Expansion platforms, respectively.

### Fine mapping

We used statistical fine-mapping as implemented in the ‘sum of single effects’ model (SuSiE) (82) using individual level genotype and protein data to identify credible sets at protein encoding loci (±500kb). Briefly, SuSiE employs a Bayesian framework for variable selection in a multiple regression problem with the aim to identify sets of independent variants each of which likely contain the true causally underlying genetic variant (82). We implemented the workflow using the R package *susieR* (v.0.11.92) and default prior and parameter settings. However, we noticed that SuSiE sometimes reports overlapping credible sets or credible sets that contained variants in high LD with already selected ones. Therefore, we adopted a grid search by first iterating the maximum number of credible sets from 2 to 10 (*L* in SuSiE terminology) and subsequently selecting the output for the maximum *L* so that none of the credible sets reported variants in LD (r^2^>0.1). We further tested for independent effects of all lead credible set variants (selecting using highest posterior inclusion probability) by including them in a joint regression model. We only report credible sets which showed genome-wide significance (p<5×10^−8^) in those joint models. We used R v.4.1 to compute regression models.

### Region-based association testing

To complement fine-mapping analysis, we computed regional association statistics at protein coding loci (±500kb) using fastGWA software provided by GCTA (v. 1.93.2beta) (83). To account for the different selection designs of the sub- and the case-cohort, we performed these analyses within each cohort separately and combined in an inverse-variance fixed-effects meta-analysis in METAL (84).

### Gene, Variant, and Protein annotation

We obtained conservation scores for all protein coding genes from gnomAD. We used the Variant Effect Predictor software (22) (version 98.3) with the --pick option to annotate all independent lead variants and proxies (r^2^>0.6) of identified pQTLs in our data set and report possible functional consequences. We collapsed pQTLs mapping to the same functional variant to reduce redundancy. We further obtained protein characteristics, e.g., glycosylation sites, from UniProt (85).

### Annotation of GWAS catalog loci

We downloaded genome-wide significant summary statistics from the GWAS catalog (date 23/03/2022; (1)) and tested whether any of the lead credible set variants (cis-pQTLs) or proxies (r^2^>0.8) have been reported to be associated with any non-proteomic trait, that is omitting any results that related to multiplex proteomic assays. Out of 347,165 entries (n=9,997 unique traits), 212,628 entries (n=5,391 unique traits) passed this and additional filtering steps (missing effect estimates, missing risk allele, and not passing genome-wide significance). For each cis-pQTL – GWAS variant mapping, we compared the reported or mapped gene (closest gene assigned by the GWAS catalog) to the protein-encoding gene at the locus.

### Phenome-wide analyses at protein-encoding loci

We performed phenome-wide analyses using statistical colocalization for 914 protein targets where we had evidence for at least one cis-pQTL. To this end, we queried the Open GWAS database (86, 87) using a defined region (±500 kb) around the protein-encoding gene body and tested whether any of the traits in the databases showed a high posterior probability (PP) of shared genetic signal with plasma concentrations of the encoded protein target using statistical colocalization (88). We chose a cut-off of PP>80% to declare that a protein target and a phenotypic trait are highly likely to share a genetic signal at a locus. We used a conservative prior setting with p_12_=1×10^−6^ and further ensured that regional sentinel variants were in strong LD (r^2^>0.8). To avoid spurious colocalization results due to imperfect overlap of SNPs, we filter all results for which the strongest cis-pQTL or sufficient proxy (r^2^>0.8) in the overlapping set was not included in the overlapping set of SNPs or if less than 500 SNPs were overlapping. We used the *igraph* package in R to visualize protein – disease colocalization results as a network to account for cross-disease dependencies established by proteins.

### Incorporation of gene expression data

We systematically tested for a shared genetic signal between plasma abundances of a protein and gene expression levels (eQTL) of the protein coding gene in 49 tissues from the GTEx project (v8) (89). We used a similar colocalization framework as described above but adopting a less stringent p_12_ prior (p_12_=1×10^−5^) to account for the higher biological prior of genetic signals in the protein encoding region. All GTEx variant-gene cis-eQTL and cis-sQTL associations from each tissue were downloaded in January 2020 from https://console.cloud.google.com/storage/browser/gtex-resources.

### Phenotypic convergence between pQTL colocalization and rare loss of function gene-burden associations

To compare the phenotypic convergence of rare loss of function gene-burden and cis-pQTLs colocalization results, we downloaded single variant and gene-burden results for 3,986 phenotypic outcomes from UK Biobank respectively which were analysed by Backman et al. (2021) (downloaded on: 07/12/2021); (64). We filtered the results for 2,939 protein coding genes covered by the Olink Explore 1536 and Explore Expansion platforms. We compared the phenotypic convergence of genes that were significant for at least one phenotypic outcome in the exome-wide association analysis at exome-wide significance (p<1×10^−6^) with the pQTLs that showed significant statistical colocalization for at least one trait (PP>80%). If ExWAS results were significant for more than one variant group for the same gene – trait association, we have filtered the results to only take forward the most significant finding.

### Multitrait colocalisation

We used hypothesis prioritisation in multi-trait colocalisation (HyPrColoc) (90) at selected protein loci to identify a shared genetic signal across various traits, including gene expression, plasma protein levels, and prioritized phenotypes from the disease-wise colocalization framework. HyPrColoc provides for each cluster three different types of output: 1) a posterior probability (PP) that all phenotypes in the cluster share a common genetic signal, 2) a regional association probability, that it, that all the phenotypes share an association with one or more variants in the region, and 3) the proportion of the PP explained by the candidate variant. We considered a highly likely alignment of a genetic signal across various phenotypes if the PP>80% and report obtained PPs otherwise.

## Data Availability

The EPIC-Norfolk data can be requested by bona fide researchers for specified scientific purposes via the study website (https://www.mrc-epid.cam.ac.uk/research/studies/epic-norfolk/). Data will either be shared through an institutional data sharing agreement or arrangements will be made for analyses to be conducted remotely without the need for data transfer.
Fine-mapped summary statistics for protein coding regions will be shared publicly following publication.
Genome-wide association studies for anthropometric phenotypes have been conducted using the UK Biobank resource (application no. 44448). Access to the UK Biobank genotype and phenotype data is open to all approved health researchers (http://www.ukbiobank.ac.uk/).

## Acknowledgements

The EPIC-Norfolk study (DOI 10.22025/2019.10.105.00004) has received funding from the Medical Research Council (MR/N003284/1 MC-UU_12015/1 and MC_UU_00006/1) and Cancer Research UK (C864/A14136). The genetics work in the EPIC-Norfolk study was funded by the Medical Research Council (MC_PC_13048). We are grateful to all the participants who have been part of the project and to the many members of the study teams at the University of Cambridge including the EPIC-Norfolk investigators, the Study Co-ordination team, the Epidemiology Field, Data and Laboratory teams who have enabled this research. Proteomics measurements were supported by a collaboration agreement between the University of Cambridge and Olink. We thank Philippa Pettingill, Ida Grundberg and Janet Kenyon for their support with quality control on the proteomic data. M.K. is supported by Gates Cambridge Trust. JCZS is supported by a 4-year Wellcome Trust PhD Studentship and the Cambridge Trust. C.L., E.W., M.P., N.K. and N.J.W. are funded by the Medical Research Council (MC_UU_00006/1). The authors thank Million Veteran Program (MVP) staff, researchers, and volunteers, who have contributed to MVP, and especially participants who previously served their country in the military and now generously agreed to enroll in the study (see https://www.research.va.gov/mvp/ for more details). We thank Friedemann Paul, Aroon Hingorani and Siamon Gordon for sharing their expertise on disease-specific examples.

## Author contributions

MK, MP and CL designed the analysis and drafted the manuscript. MK, MP and EW have performed the bioinformatics analyses. JCZS and NK have performed the quality control and data preparation of the proteomic data. SL contributed to the interpretation and curation of disease examples. NJW is PI of the EPIC-Norfolk study. All authors contributed to the interpretation of the results and critically reviewed the manuscript.

## Data availability

The EPIC-Norfolk data can be requested by bona fide researchers for specified scientific purposes via the study website (https://www.mrc-epid.cam.ac.uk/research/studies/epic-norfolk/). Data will either be shared through an institutional data sharing agreement or arrangements will be made for analyses to be conducted remotely without the need for data transfer.

Fine-mapped summary statistics for protein coding regions will be shared publicly following publication.

Genome-wide association studies for anthropometric phenotypes have been conducted using the UK Biobank resource (application no. 44448). Access to the UK Biobank genotype and phenotype data is open to all approved health researchers (http://www.ukbiobank.ac.uk/).

## References

1. Buniello A, MacArthur JAL, Cerezo M, Harris LW, Hayhurst J, Malangone C, et al. The NHGRI-EBI GWAS Catalog of published genome-wide association studies, targeted arrays and summary statistics 2019. Nucleic Acids Res. 2019;47(D1):D1005–D12.

2. Barbeira AN, Bonazzola R, Gamazon ER, Liang Y, Park Y, Kim-Hellmuth S, et al. Exploiting the GTEx resources to decipher the mechanisms at GWAS loci. Genome Biol. 2021;22(1):49.

3. Moore JE, Purcaro MJ, Pratt HE, Epstein CB, Shoresh N, Adrian J, et al. Expanded encyclopaedias of DNA elements in the human and mouse genomes. Nature. 2020;583(7818):699–710.

4. Abell NS, DeGorter MK, Gloudemans MJ, Greenwald E, Smith KS, He Z, et al. Multiple causal variants underlie genetic associations in humans. Science. 2022;375(6586):1247–54.

5. Suhre K, Arnold M, Bhagwat AM, Cotton RJ, Engelke R, Raffler J, et al. Connecting genetic risk to disease end points through the human blood plasma proteome. Nat Commun. 2017;8:14357.

6. Sun BB, Maranville JC, Peters JE, Stacey D, Staley JR, Blackshaw J, et al. Genomic atlas of the human plasma proteome. Nature. 2018;558(7708):73–9.

7. Folkersen L, Fauman E, Sabater-Lleal M, Strawbridge RJ, Frånberg M, Sennblad B, et al. Mapping of 79 loci for 83 plasma protein biomarkers in cardiovascular disease. PLoS Genet. 2017;13(4):e1006706.

8. Yao C, Chen G, Song C, Keefe J, Mendelson M, Huan T, et al. Author Correction: Genome-wide mapping of plasma protein QTLs identifies putatively causal genes and pathways for cardiovascular disease. Nat Commun. 2018;9(1):3853.

9. Gilly A, Park YC, Png G, Barysenka A, Fischer I, Bjørnland T, et al. Whole-genome sequencing analysis of the cardiometabolic proteome. Nat Commun. 2020;11(1):6336.

10. Ferkingstad E, Sulem P, Atlason BA, Sveinbjornsson G, Magnusson MI, Styrmisdottir EL, et al. Large-scale integration of the plasma proteome with genetics and disease. Nat Genet. 2021;53(12):1712–21.

11. Gudjonsson A, Gudmundsdottir V, Axelsson GT, Gudmundsson EF, Jonsson BG, Launer LJ, et al. A genome-wide association study of serum proteins reveals shared loci with common diseases. Nat Commun. 2022;13(1):480.

12. Katz DH, Tahir UA, Bick AG, Pampana A, Ngo D, Benson MD, et al. Whole Genome Sequence Analysis of the Plasma Proteome in Black Adults Provides Novel Insights Into Cardiovascular Disease. Circulation. 2022;145(5):357–70.

13. Png G, Barysenka A, Repetto L, Navarro P, Shen X, Pietzner M, et al. Mapping the serum proteome to neurological diseases using whole genome sequencing. Nat Commun. 2021;12(1):7042.

14. Pietzner M, Wheeler E, Carrasco-Zanini J, Kerrison ND, Oerton E, Koprulu M, et al. Synergistic insights into human health from aptamer- and antibody-based proteomic profiling. Nat Commun. 2021;12(1):6822.

15. Pietzner M, Wheeler E, Carrasco-Zanini J, Cortes A, Koprulu M, Wörheide MA, et al. Mapping the proteo-genomic convergence of human diseases. Science. 2021;374(6569):eabj1541.

16. Zhang J, Dutta D, Köttgen A, Tin A, Schlosser P, Grams ME, et al. Plasma proteome analyses in individuals of European and African ancestry identify cis-pQTLs and models for proteome-wide association studies. Nat Genet. 2022;54(5):593–602.

17. Sun BB, Chiou J, Traylor M, Benner C, Hsu Y-H, Richardson TG, et al. Genetic regulation of the human plasma proteome in 54,306 UK Biobank participants. bioRxiv. 2022:2022.06.17.496443.

18. Folkersen L, Gustafsson S, Wang Q, Hansen DH, Hedman Å, Schork A, et al. Genomic and drug target evaluation of 90 cardiovascular proteins in 30,931 individuals. Nat Metab. 2020;2(10):1135–48.

19. Enroth S, Johansson A, Enroth SB, Gyllensten U. Strong effects of genetic and lifestyle factors on biomarker variation and use of personalized cutoffs. Nat Commun. 2014;5:4684.

20. Wang G, Sarkar A, Carbonetto P, Stephens M. A simple new approach to variable selection in regression, with application to genetic fine mapping. Journal of the Royal Statistical Society: Series B (Statistical Methodology). 2020;82(5):1273–300.

21. Day N, Oakes S, Luben R, Khaw KT, Bingham S, Welch A, et al. EPIC-Norfolk: study design and characteristics of the cohort. European Prospective Investigation of Cancer. Br J Cancer. 1999;80 Suppl 1:95–103.

22. McLaren W, Gil L, Hunt SE, Riat HS, Ritchie GR, Thormann A, et al. The Ensembl Variant Effect Predictor. Genome Biol. 2016;17(1):122.

23. Vujkovic M, Keaton JM, Lynch JA, Miller DR, Zhou J, Tcheandjieu C, et al. Discovery of 318 new risk loci for type 2 diabetes and related vascular outcomes among 1.4 million participants in a multi-ancestry meta-analysis. Nat Genet. 2020;52(7):680–91.

24. Spracklen CN, Horikoshi M, Kim YJ, Lin K, Bragg F, Moon S, et al. Identification of type 2 diabetes loci in 433,540 East Asian individuals. Nature. 2020;582(7811):240–5.

25. Mahajan A, Spracklen CN, Zhang W, Ng MCY, Petty LE, Kitajima H, et al. Multi-ancestry genetic study of type 2 diabetes highlights the power of diverse populations for discovery and translation. Nat Genet. 2022;54(5):560–72.

26. McDonald TJ, Nilsson G, Vagne M, Ghatei M, Bloom SR, Mutt V. A gastrin releasing peptide from the porcine nonantral gastric tissue. Gut. 1978;19(9):767–74.

27. McDonald TJ, Jörnvall H, Nilsson G, Vagne M, Ghatei M, Bloom SR, et al. Characterization of a gastrin releasing peptide from porcine non-antral gastric tissue. Biochem Biophys Res Commun. 1979;90(1):227–33.

28. Ladenheim EE, Taylor JE, Coy DH, Moore KA, Moran TH. Hindbrain GRP receptor blockade antagonizes feeding suppression by peripherally administered GRP. Am J Physiol. 1996;271(1 Pt 2):R180–4.

29. Ladenheim EE, Hampton LL, Whitney AC, White WO, Battey JF, Moran TH. Disruptions in feeding and body weight control in gastrin-releasing peptide receptor deficient mice. J Endocrinol. 2002;174(2):273–81.

30. Persson K, Gingerich RL, Nayak S, Wada K, Wada E, Ahrén B. Reduced GLP-1 and insulin responses and glucose intolerance after gastric glucose in GRP receptor-deleted mice. Am J Physiol Endocrinol Metab. 2000;279(5):E956–62.

31. Gutzwiller JP, Drewe J, Hildebrand P, Rossi L, Lauper JZ, Beglinger C. Effect of intravenous human gastrin-releasing peptide on food intake in humans. Gastroenterology. 1994;106(5):1168– 73.

32. Mhalhal TR, Washington MC, Newman KD, Heath JC, Sayegh AI. Combined gastrin releasing peptide-29 and glucagon like peptide-1 reduce body weight more than each individual peptide in diet-induced obese male rats. Neuropeptides. 2018;67:71–8.

33. Loh PR, Tucker G, Bulik-Sullivan BK, Vilhjálmsson BJ, Finucane HK, Salem RM, et al. Efficient Bayesian mixed-model analysis increases association power in large cohorts. Nat Genet. 2015;47(3):284–90.

34. Gaziano JM, Concato J, Brophy M, Fiore L, Pyarajan S, Breeling J, et al. Million Veteran Program: A mega-biobank to study genetic influences on health and disease. J Clin Epidemiol. 2016;70:214–23.

35. Pulit SL, Stoneman C, Morris AP, Wood AR, Glastonbury CA, Tyrrell J, et al. Meta-analysis of genome-wide association studies for body fat distribution in 694 649 individuals of European ancestry. Hum Mol Genet. 2019;28(1):166–74.

36. Frullanti E, Berking C, Harbeck N, Jézéquel P, Haugen A, Mawrin C, et al. Meta and pooled analyses of FGFR4 Gly388Arg polymorphism as a cancer prognostic factor. Eur J Cancer Prev. 2011;20(4):340–7.

37. Chou CH, Hsieh MJ, Chuang CY, Lin JT, Yeh CM, Tseng PY, et al. Functional FGFR4 Gly388Arg polymorphism contributes to oral squamous cell carcinoma susceptibility. Oncotarget. 2017;8(56):96225–38.

38. Xiong SW, Ma J, Feng F, Fu W, Shu SR, Ma T, et al. Functional FGFR4 Gly388Arg polymorphism contributes to cancer susceptibility: Evidence from meta-analysis. Oncotarget. 2017;8(15):25300–9.

39. Ulaganathan VK, Sperl B, Rapp UR, Ullrich A. Germline variant FGFR4 p.G388R exposes a membrane-proximal STAT3 binding site. Nature. 2015;528(7583):570–4.

40. Shin DJ, Osborne TF. FGF15/FGFR4 integrates growth factor signaling with hepatic bile acid metabolism and insulin action. J Biol Chem. 2009;284(17):11110–20.

41. Ge H, Zhang J, Gong Y, Gupte J, Ye J, Weiszmann J, et al. Fibroblast growth factor receptor 4 (FGFR4) deficiency improves insulin resistance and glucose metabolism under diet-induced obesity conditions. J Biol Chem. 2014;289(44):30470–80.

42. Wu X, Ge H, Lemon B, Weiszmann J, Gupte J, Hawkins N, et al. Selective activation of FGFR4 by an FGF19 variant does not improve glucose metabolism in ob/ob mice. Proc Natl Acad Sci U S A. 2009;106(34):14379–84.

43. Huang X, Yang C, Luo Y, Jin C, Wang F, McKeehan WL. FGFR4 prevents hyperlipidemia and insulin resistance but underlies high-fat diet induced fatty liver. Diabetes. 2007;56(10):2501–10.

44. Lutz SZ, Hennige AM, Peter A, Kovarova M, Totsikas C, Machann J, et al. The Gly385(388)Arg Polymorphism of the FGFR4 Receptor Regulates Hepatic Lipogenesis Under Healthy Diet. J Clin Endocrinol Metab. 2019;104(6):2041–53.

45. Micha R, Khatibzadeh S, Shi P, Fahimi S, Lim S, Andrews KG, et al. Global, regional, and national consumption levels of dietary fats and oils in 1990 and 2010: a systematic analysis including 266 country-specific nutrition surveys. BMJ. 2014;348:g2272.

46. Lappalainen T, MacArthur DG. From variant to function in human disease genetics. Science. 2021;373(6562):1464–8.

47. Filippi M, Bar-Or A, Piehl F, Preziosa P, Solari A, Vukusic S, et al. Multiple sclerosis. Nat Rev Dis Primers. 2018;4(1):43.

48. Consortium IMSG. Multiple sclerosis genomic map implicates peripheral immune cells and microglia in susceptibility. Science. 2019;365(6460).

49. Yin X, Kim K, Suetsugu H, Bang SY, Wen L, Koido M, et al. Meta-analysis of 208370 East Asians identifies 113 susceptibility loci for systemic lupus erythematosus. Ann Rheum Dis. 2021;80(5):632–40.

50. Kaneko KJ, Kohn MJ, Liu C, DePamphilis ML. The acrosomal protein Dickkopf-like 1 (DKKL1) is not essential for fertility. Fertil Steril. 2010;93(5):1526–32.

51. Uhlen M, Karlsson MJ, Zhong W, Tebani A, Pou C, Mikes J, et al. A genome-wide transcriptomic analysis of protein-coding genes in human blood cells. Science. 2019;366(6472).

52. Orrù V, Steri M, Sidore C, Marongiu M, Serra V, Olla S, et al. Complex genetic signatures in immune cells underlie autoimmunity and inform therapy. Nat Genet. 2020;52(10):1036–45.

53. Cencioni MT, Mattoscio M, Magliozzi R, Bar-Or A, Muraro PA. B cells in multiple sclerosis - from targeted depletion to immune reconstitution therapies. Nat Rev Neurol. 2021;17(7):399– 414.

54. Granqvist M, Boremalm M, Poorghobad A, Svenningsson A, Salzer J, Frisell T, et al. Comparative Effectiveness of Rituximab and Other Initial Treatment Choices for Multiple Sclerosis. JAMA Neurol. 2018;75(3):320–7.

55. de Rojas I, Moreno-Grau S, Tesi N, Grenier-Boley B, Andrade V, Jansen IE, et al. Common variants in Alzheimer’s disease and risk stratification by polygenic risk scores. Nat Commun. 2021;12(1):3417.

56. Richardson TG, Sanderson E, Elsworth B, Tilling K, Davey Smith G. Use of genetic variation to separate the effects of early and later life adiposity on disease risk: mendelian randomisation study. BMJ. 2020;369:m1203.

57. Liu H, Leo C, Chen X, Wong BR, Williams LT, Lin H, et al. The mechanism of shared but distinct CSF-1R signaling by the non-homologous cytokines IL-34 and CSF-1. Biochim Biophys Acta. 2012;1824(7):938–45.

58. Delaney C, Farrell M, Doherty CP, Brennan K, O’Keeffe E, Greene C, et al. Attenuated CSF-1R signalling drives cerebrovascular pathology. EMBO Mol Med. 2021;13(2):e12889.

59. Morris JA, Kemp JP, Youlten SE, Laurent L, Logan JG, Chai RC, et al. An atlas of genetic influences on osteoporosis in humans and mice. Nat Genet. 2019;51(2):258–66.

60. Lotta LA, Wittemans LBL, Zuber V, Stewart ID, Sharp SJ, Luan J, et al. Association of Genetic Variants Related to Gluteofemoral vs Abdominal Fat Distribution With Type 2 Diabetes, Coronary Disease, and Cardiovascular Risk Factors. JAMA. 2018;320(24):2553–63.

61. Acosta-Herrera M, Kerick M, González-Serna D, Wijmenga C, Franke A, Gregersen PK, et al. Genome-wide meta-analysis reveals shared new loci in systemic seropositive rheumatic diseases. Ann Rheum Dis. 2019;78(3):311–9.

62. Robertson CC, Inshaw JRJ, Onengut-Gumuscu S, Chen WM, Santa Cruz DF, Yang H, et al. Fine-mapping, trans-ancestral and genomic analyses identify causal variants, cells, genes and drug targets for type 1 diabetes. Nat Genet. 2021;53(7):962–71.

63. Clarke LA, Giugliani R, Guffon N, Jones SA, Keenan HA, Munoz-Rojas MV, et al. Genotype-phenotype relationships in mucopolysaccharidosis type I (MPS I): Insights from the International MPS I Registry. Clin Genet. 2019;96(4):281–9.

64. Backman JD, Li AH, Marcketta A, Sun D, Mbatchou J, Kessler MD, et al. Exome sequencing and analysis of 454,787 UK Biobank participants. Nature. 2021;599(7886):628–34.

65. Szustakowski JD, Balasubramanian S, Kvikstad E, Khalid S, Bronson PG, Sasson A, et al. Advancing human genetics research and drug discovery through exome sequencing of the UK Biobank. Nat Genet. 2021;53(7):942–8.

66. Plenge RM, Scolnick EM, Altshuler D. Validating therapeutic targets through human genetics. Nat Rev Drug Discov. 2013;12(8):581–94.

67. Kathiresan S, Willer CJ, Peloso GM, Demissie S, Musunuru K, Schadt EE, et al. Common variants at 30 loci contribute to polygenic dyslipidemia. Nat Genet. 2009;41(1):56–65.

68. Lemke G. How macrophages deal with death. Nat Rev Immunol. 2019;19(9):539–49.

69. Miyanishi M, Tada K, Koike M, Uchiyama Y, Kitamura T, Nagata S. Identification of Tim4 as a phosphatidylserine receptor. Nature. 2007;450(7168):435–9.

70. Kuchroo VK, Dardalhon V, Xiao S, Anderson AC. New roles for TIM family members in immune regulation. Nat Rev Immunol. 2008;8(8):577–80.

71. Mouse Genome Database (MGD) at the Mouse Genome Informatics website, The Jackson Laboratory 2022 [Available from: http://www.informatics.jax.org/.

72. Fazio S, Hasty AH, Carter KJ, Murray AB, Price JO, Linton MF. Leukocyte low density lipoprotein receptor (LDL-R) does not contribute to LDL clearance in vivo: bone marrow transplantation studies in the mouse. J Lipid Res. 1997;38(2):391–400.

73. Magalhaes MS, Smith P, Portman JR, Jackson-Jones LH, Bain CC, Ramachandran P, et al. Role of Tim4 in the regulation of ABCA1. Nat Commun. 2021;12(1):4434.

74. Koelwyn GJ, Corr EM, Erbay E, Moore KJ. Regulation of macrophage immunometabolism in atherosclerosis. Nat Immunol. 2018;19(6):526–37.

75. Foks AC, Engelbertsen D, Kuperwaser F, Alberts-Grill N, Gonen A, Witztum JL, et al. Blockade of Tim-1 and Tim-4 Enhances Atherosclerosis in Low-Density Lipoprotein Receptor-Deficient Mice. Arterioscler Thromb Vasc Biol. 2016;36(3):456–65.

76. Alsheikh AJ, Wollenhaupt S, King EA, Reeb J, Ghosh S, Stolzenburg LR, et al. The landscape of GWAS validation; systematic review identifying 309 validated non-coding variants across 130 human diseases. BMC Med Genomics. 2022;15(1):74.

77. Gudmundsson S, Karczewski KJ, Francioli LC, Tiao G, Cummings BB, Alföldi J, et al. Addendum: The mutational constraint spectrum quantified from variation in 141,456 humans. Nature. 2021;597(7874):E3–E4.

78. Mostafavi H, Spence JP, Naqvi S, Pritchard JK. Limited overlap of eQTLs and GWAS hits due to systematic differences in discovery. bioRxiv. 2022:2022.05.07.491045.

79. Akbari P, Gilani A, Sosina O, Kosmicki JA, Khrimian L, Fang YY, et al. Sequencing of 640,000 exomes identifies GPR75 variants associated with protection from obesity. Science. 2021;373(6550).

80. Zhong W, Edfors F, Gummesson A, Bergström G, Fagerberg L, Uhlén M. Next generation plasma proteome profiling to monitor health and disease. Nat Commun. 2021;12(1):2493.

81. Assarsson E, Lundberg M, Holmquist G, Björkesten J, Thorsen SB, Ekman D, et al. Homogenous 96-plex PEA immunoassay exhibiting high sensitivity, specificity, and excellent scalability. PLoS One. 2014;9(4):e95192.

82. Wang G, Sarkar A, Car bonetto p, S tephens M. A simple new approach to variable selection in regression, with application to genetic fine mapping. Royal Statistical Society; 2020. p. 1273–300.

83. Jiang L, Zheng Z, Qi T, Kemper KE, Wray NR, Visscher PM, et al. A resource-efficient tool for mixed model association analysis of large-scale data. Nat Genet. 2019;51(12):1749–55.

84. Willer CJ, Li Y, Abecasis GR. METAL: fast and efficient meta-analysis of genomewide association scans. Bioinformatics. 2010;26(17):2190–1.

85. Consortium U. UniProt: the universal protein knowledgebase in 2021. Nucleic Acids Res. 2021;49(D1):D480–D9.

86. Elsworth B, Lyon M, Alexander T, Liu Y, Matthews P, Hallett J, et al. The MRC IEU OpenGWAS data infrastructure. bioRxiv. 2020:2020.08.10.244293.

87. Hemani G, Zheng J, Elsworth B, Wade KH, Haberland V, Baird D, et al. The MR-Base platform supports systematic causal inference across the human phenome. Elife. 2018;7.

88. Giambartolomei C, Vukcevic D, Schadt EE, Franke L, Hingorani AD, Wallace C, et al. Bayesian test for colocalisation between pairs of genetic association studies using summary statistics. PLoS Genet. 2014;10(5):e1004383.

89. Consortium G. The GTEx Consortium atlas of genetic regulatory effects across human tissues. Science. 2020;369(6509):1318–30.

90. Foley CN, Staley JR, Breen PG, Sun BB, Kirk PDW, Burgess S, et al. A fast and efficient colocalization algorithm for identifying shared genetic risk factors across multiple traits. Nat Commun. 2021;12(1):764.

