## Supplementary Materials for "From genome to phenome via the proteome: broad capture, antibody-based proteomics to explore disease mechanisms"

### Supplementary Figures

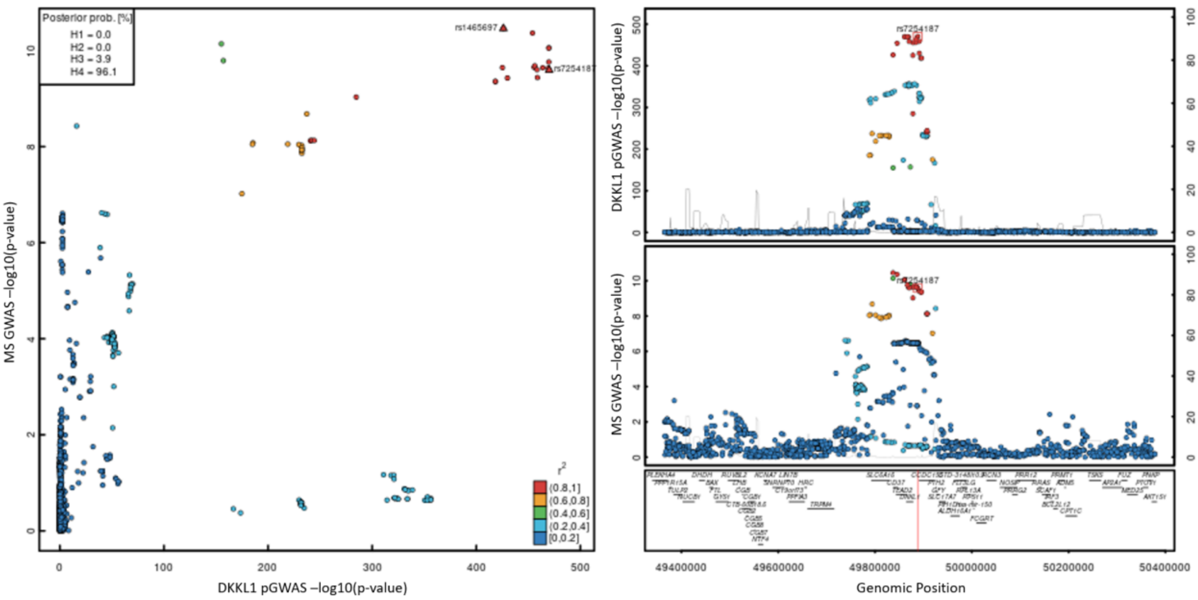


**Supplementary Figure 1: Locusplot comparing association statistics for DDKL1 and multiple sclerosis (MS).** The left panel displays a comparison of -$\log_{10}$-transformed p-values from GWAS summary statistics for genetic variants in a 500 kb region around the lead signal on chromosome 19. Colouring was done based on linkage disequilibrium with the lead variant for the protein. The right panel is a stacked locuszoom plot (DKKL1 on top, MS on the bottom) with annotation of protein-encoding genes underneath. Location of the lead variant is indicated by a red line.

### Supplementary Table titles

**Supplementary Table 1. Demographics of the European Prospective Investigation into Cancer (EPIC)-Norfolk study.**

**Supplementary Table 2. Independent credible sets for protein targets at protein encoding loci (±500kb) identified by fine-mapping.** This table includes all 1,553 independent credible sets for protein targets, annotated by the lead variant in each credible set and their functional annotations.

**Supplementary Table 3. Results from colocalization analysis with gene expression trait loci (eQTLs) from the GTEx version 8 release at the protein encoding loci (±500kb).** The table contains all gene expression - protein pairings where the posterior probability of a shared signal was above 80%. For each protein target where we have observed strong statistical colocalization (posterior probability of shared signal > 80%), all of the significant tissues are listed, and relevant posterior probabilities are provided in brackets.

**Supplementary Table 4. Protein-target – phenotype pairs with strong evidence of colocalization at the protein encoding locus (±500kb).** The table contains all protein – phenotype connections where the posterior probability of shared signal was above 80% and regional sentinel variants were in strong linkage disequilibrium ($r^2$>0.8). Information on draggability based on common gene entries from Finan et al. (2017) (1).

**Supplementary Table 5. pQTL mapping of candidate causal genes at previously reported GWAS loci from GWAS Catalog.** For each mapping cis-pQTL – GWAS variant pair all curated traits are listed, and two columns indicate whether the protein-encoding genes has been reported as the causal gene at this locus or is the closest gene at this locus.

**Supplementary Table 6. Phenotypic convergence between pQTL colocalization and rare loss of function gene-burden associations.** This table includes information on the phenotypic convergence between pQTL colocalization and rare loss of function gene-burden associations from Backman et al (2021) (2) for 40 overlapping genes. Following manual review to harmonize phenotype definitions, converge flag column indicates the phenotypic convergence amongst the list of phenotypes for 40 overlapping genes. Information on draggability based on common gene entries from Finan et al. (2017) (1).

### References


1. Finan C, Gaulton A, Kruger FA, Lumbers RT, Shah T, Engmann J, et al. The druggable genome and support for target identification and validation in drug development. Sci Transl Med. 2017;9(383).
2. Backman JD, Li AH, Marcketta A, Sun D, Mbatchou J, Kessler MD, et al. Exome sequencing and analysis of 454,787 UK Biobank participants. Nature. 2021;599(7886):628-34.
